# Population Genomics of *Salmonella* Enteritidis in Saudi Arabia Reveals Globally Circulating Food- and Human-Associated Lineages and Plasmid-Mediated Antimicrobial Resistance

**DOI:** 10.64898/2026.09.03.26362153

**Authors:** Fahad Alreshoodi, Ge Zhou, Jiayi Huang, Natalie Hinkova, Amani Alsufyani, Norah M. Alotaibi, Mohammed S. Alarawi, Khaloud O. Alzahrani, Shahad A. Al Salman, Manal A. Almusa, Abdullah A. Alajlan, Elaf A. Alshdokhi, Afnan Althobaiti, Hatim H. Almutairi, Saleh I. Al-Akeel, Anfal Al-Ajmi, Abdulmohsen L. AlHarbi, Malfi S. Al Rashidy, Sara Iftikhar, Omniya Fallatah, Manuel Banzhaf, Suliman Alajel, Abiola Senok, Lenah Mukhtar, Séamus Fanning, Danesh Moradigaravand

## Abstract

**Background:** *Salmonella enterica* serovar Enteritidis (*S.* Enteritidis) is one of the leading causes of foodborne gastroenteritis worldwide and an increasingly important driver of antimicrobial-resistant infections. Despite its public health importance, genomic surveillance of *S.* Enteritidis remains sparse across the Middle East, where integrated One Health studies linking human, food, and animal reservoirs are limited. This lack of regional genomic data has hindered understanding of the population structure, transmission dynamics, and dissemination of antimicrobial-resistant lineages. We therefore performed a large-scale One Health genomic investigation of *S.* Enteritidis in the Kingdom of Saudi Arabia, integrating human, food, poultry, and global genomic datasets to characterize its evolutionary history, transmission dynamics, and antimicrobial resistance.

**Methods:** A total of 220 Salmonella enterica serovar Enteritidis (S. Enteritidis) isolates obtained in Saudi Arabia from human clinical infections and food products between 2020 and 2023 underwent whole-genome sequencing. To position these isolates within a global evolutionary context, they were integrated with all publicly available S. Enteritidis genomes, yielding a comparative dataset of 397 genomes. A comprehensive genomic analysis framework was employed, encompassing population genomic, phylogenetic, and phylodynamic analyses to define population structure, infer evolutionary history, and investigate transmission dynamics. In addition, the genomic determinants of antimicrobial resistance and virulence were characterized, with particular emphasis on plasmid architecture.

**Results:** Isolates from Saudi Arabia belonged predominantly to a single globally distributed epidemiological cluster of *S.* Enteritidis and were embedded within globally circulating transmission networks rather than forming a distinct endemic population. Within this cluster, the Saudi isolates were distributed across seven Bayesian Analysis of Population Structure (BAPS) groups, representing distinct genomic sub-clones. Multiple lineages contained closely related isolates from Europe, North America, Oceania, and the Middle East, indicating repeated international introductions and extensive global connectivity. Human and food isolates were highly intermixed throughout the phylogeny, with food-associated isolates frequently occupying central positions within transmission networks, consistent with food reservoirs acting as dissemination hubs linking local and international transmission. Phylodynamic analyses indicated that the dominant Saudi-associated lineages emerged within the past 10–15 years and underwent rapid demographic expansion following introduction into the country. Recently emerged epidemic lineages carried significantly higher antimicrobial resistance burdens than ancestral populations and were characterized by near fixation of *bla*_TEM-1_ and *tetA* on IncX1 plasmids, whereas clinically important ESBL genes (*bla*_CTX-M-8_ and *bla*_SHV-12_) were detected exclusively in human-associated isolates on hybrid resistance–virulence plasmids.

**Conclusions:** The population of *S.* Enteritidis in Saudi Arabia is shaped by repeated introductions of globally circulating clones, transmission between food and human reservoirs, and the dissemination of resistance plasmids across lineages. The emergence of successful epidemic clones is linked to plasmid-mediated acquisition of antimicrobial resistance determinants. These findings emphasize the importance of integrated One Health surveillance strategies for controlling foodborne transmission and monitoring the emergence of antimicrobial-resistant epidemic lineages.

## Introduction

Foodborne diseases remain a major global public health burden, with non-typhoidal *Salmonella* among the leading bacterial causes of illness worldwide (1, 2). Within this group, *Salmonella enterica* subsp. *enterica* serovar Enteritidis (*S.* Enteritidis) is a predominant cause of foodborne infection, contributing substantially to morbidity, economic losses, and healthcare costs across both high- and low-income settings (3–6). In the European Union, more than 60,000 salmonellosis cases were reported in 2021, with *S.* Enteritidis accounting for the largest proportion of reported infections (7). A similar epidemiological pattern is observed in North America, where *S.* Enteritidis remains one of the most frequently reported serovars associated with human infections and foodborne outbreaks (8). Across the Middle East and North Africa, *S.* Enteritidis and *S.* Typhimurium are the predominant circulating serovars (6, 9).

*S.* Enteritidis has long been associated with poultry production and poultry-derived foods, particularly eggs and chicken meat. Since the global expansion of *S.* Enteritidis in the 1980s (10), driven by widespread contamination of poultry products, population genomic studies have shown that the majority of global isolates belong to the ST11 lineage (11), which is widely distributed across human, animal, and food-associated reservoirs (3, 12–19). Within this globally disseminated lineage, antimicrobial resistance (AMR) has increasingly been reported, including multidrug-resistant (MDR) and fluoroquinolone-resistant sub-lineages (5, 12, 13, 20–22). The emergence and expansion of resistance within this lineage are likely due to persistent selective pressure driven by sustained antimicrobial use in poultry and other animal production systems, for treatment, disease prevention and growth promotion (23–25).

Whole genome sequencing has substantially improved the resolution of *S.* Enteritidis surveillance, enabling the differentiation of closely related isolates within the globally dominant *S.* Enteritidis ST11 and strengthening source attribution between human and poultry-associated isolates (3, 4, 26). Genomic studies have demonstrated that this lineage is genetically heterogeneous, consisting of multiple phylogenetically distinct sub-lineages with geographic and ecological associations. These sub-lineages differ in accessory genome composition, including plasmid content and antimicrobial resistance determinants, highlighting the role of horizontal gene transfer and local selective pressures in shaping the evolution of this globally disseminated lineage (12, 13, 20, 26). Furthermore, large-scale genomic studies suggest that the global spread of *S.* Enteritidis ST11 has been facilitated by centralized poultry breeding systems and transnational trade networks, while regional diversification has contributed to the emergence of distinct epidemiological and resistance profiles (3, 5). Previous genomic studies have also highlighted the central role of plasmids in the evolution of *S.* Enteritidis, serving as major vehicles for the acquisition and dissemination of AMR and virulence determinants. The conserved IncFIB(S)/IncFII(S) virulence plasmid is a hallmark of the globally dominant ST11 lineage, while additional plasmid types, particularly IncX1, have been implicated in the spread of acquired AMR (27, 28). These mobile genetic elements frequently carry genes conferring resistance to β-lactams (*bla*_TEM_), tetracyclines (*tetA*), sulfonamides (*sul*), trimethoprim (*dfrA* family), and aminoglycosides (*aph*(*3*) and *aph*(*6*)) (5, 16, 17, 29), underscoring the pivotal role of horizontal gene transfer in shaping the resistance landscape of this globally disseminated lineage (24). More recently, the emergence of clinically important extended-spectrum β-lactamase (ESBL) genes, including *bla*_CTX-M-8_, has further demonstrated the capacity of ST11 to expand its resistance repertoire through plasmid-mediated gene acquisition (30–32).

Despite these advances and national-level studies (15–17, 19, 20, 22, 26), comprehensive genomic investigations of *S.* Enteritidis from diverse human and food sources in West Asia remain limited, hindering a thorough understanding of the local population structure and the regional epidemiology of MDR ST11 lineages (6, 9). As a major hub for international travel, food trade, and poultry production, West Asia is likely to play an important role in the introduction and dissemination of globally circulating *S.* Enteritidis lineages, yet the genomic processes underpinning these dynamics remain poorly understood. Within this broader regional context, the Kingdom of Saudi Arabia (KSA) provides an especially informative setting in which to investigate these processes, given its integration into regional and global food supply networks, high poultry consumption (33), complex and highly mobile population structure, and its role as a major hub for international mass gatherings (34). Together, extensive food importation and large-scale human mobility create multiple potential routes for the introduction, transmission, and onward dissemination of *S.* Enteritidis lineages.

Here, we conducted a large-scale population genomic study of *S.* Enteritidis ST11 isolated from human clinical infections and food products, including retail chicken meat, across the KSA. The country’s retail chicken market is one of the largest protein sectors in the region, driven by high per-capita consumption (approximately 40–50 kg per person annually) and strong demand from both retail and food-service sectors (33). Recent national surveillance data indicate that reported salmonellosis incidence in the KSA increased by 123.2% between 2015 and 2023, highlighting a growing public health challenge and the need for enhanced genomic surveillance and targeted control strategies. By integrating comprehensive sampling across food and clinical settings with global genomic data, we resolved the population structure of *S.* Enteritidis ST11, reconstructed transmission links between food and human infections, and identified the evolutionary and plasmid-mediated mechanisms driving the emergence and dissemination of resistant lineages on epidemiological timescales.

## Results

### National genome surveillance of human and food-associated *S*. Enteritidis in Saudi Arabia

A systematic collection of 220 S. Enteritidis ST11 isolates obtained across Saudi Arabia between 2020 and 2023 from both human and food sources was analyzed using whole-genome sequencing data. The collection comprised 97 clinical isolates obtained from human infections, predominantly stool specimens (n=72) and blood specimens (n=11), and 123 food-associated isolates obtained primarily from chicken products (n=113). Clinical isolates were collected from hospitals across multiple regions of Saudi Arabia, including the western region (Makkah), southern regions (Najran and Asir), central regions (Riyadh, Al-Jouf, and Al-Qassim), the Eastern Province, and the northern region of Tabuk. Sampling was particularly dense in Jeddah (West) and Sharurah (South) (Supplementary Table 1 and Figure S1). Food-associated isolates were primarily recovered from major population centres, including Riyadh (n=47), Dammam (n=44), and Jeddah (n=29) (Supplementary Table 1). Across the entire collection, 124/220 (56.4%) isolates were classified as MDR, 73/220 (33.2%) as non-MDR, and 23/220 (10.5%) as susceptible. ESBL-positive isolates were uncommon, accounting for only 4/220 (1.8%) isolates, including three clinical isolates and one food-associated isolate. Resistance status was significantly associated with isolate source (Pearson’s χ² = 32.32, p-value < 0.01; Fisher’s exact test, p-value < 0.01). MDR isolates were more frequent among clinical isolates (68/97, 70.1%) than food isolates (56/123, 45.5%), whereas non-MDR status was more common among food isolates (60/123, 48.8%) than clinical isolates (13/97, 13.4%), suggesting stronger selection for multidrug resistance in the clinical setting. The geographic, genomic, and source distributions showed that the collection contained broadly distributed *S.* Enteritidis isolates across Saudi Arabia from both human and food sources, with distinct antimicrobial resistance profiles between the two sources.

### Global population structure and international connectivity of Saudi Arabian *S*. Enteritidis isolates

To contextualize the *S.* Enteritidis collection under study within the global population structure of this serovar, we combined the Saudi genomes with publicly available *S.* Enteritidis ST11 genomes retrieved from the NCBI Pathogen Detection database, generating a comparative dataset of 397 genomes. This dataset included all publicly available isolates belonging to the same NCBI Pathogen Detection cluster (PDS000026888.159). Clusters are defined using wgMLST-based single-linkage clustering, whereby genomes differing by no more than 25 wgMLST alleles from at least one other genome in the cluster are grouped together (see Methods). The external collection comprised 189 clinical, 17 food-associated, and 3 environmental isolates. Bayesian Analysis of Population Structure (BAPS) identified twelve groups (BAPS-1 to BAPS-12), each comprising both Saudi and global isolates from diverse geographic regions and epidemiological sources. These groups largely corresponded to distinct clades in the core-genome phylogeny reconstructed from 1,218 core-genome SNPs (Figure 1A). Except for BAPS-5 and BAPS-10, all BAPS groups contained a mixture of clinical and food-associated isolates originating from multiple continents, consistent with the global circulation of the cluster. Likewise, isolates from human, food, and environmental sources were interspersed throughout the phylogeny rather than forming source-specific clades. Principal component analysis (PCA) was consistent with the phylogenetic reconstruction, with isolates clustering predominantly according to their BAPS assignments rather than source. No clear separation was observed among human, food, and environmental isolates (Figure 1B), further supporting the sharing of genotypes across epidemiological reservoirs. In contrast, the twelve BAPS groups formed localized but partially overlapping clusters (Figure 1C), reflecting their overall genetic relatedness. Hierarchical BAPS clustering further resolved the population into two broad genetic groups. The first comprised BAPS-1 to BAPS-8, including all Saudi isolates, and represented the dominant, closely related lineage within the dataset. The second comprised the more genetically divergent BAPS-9 to BAPS-12. Together with the phylogenetic analysis, these findings indicate that the Saudi isolates are embedded within a globally circulating *S.* Enteritidis ST11 population characterized by extensive geographic and ecological mixing.

**Figure 1.**
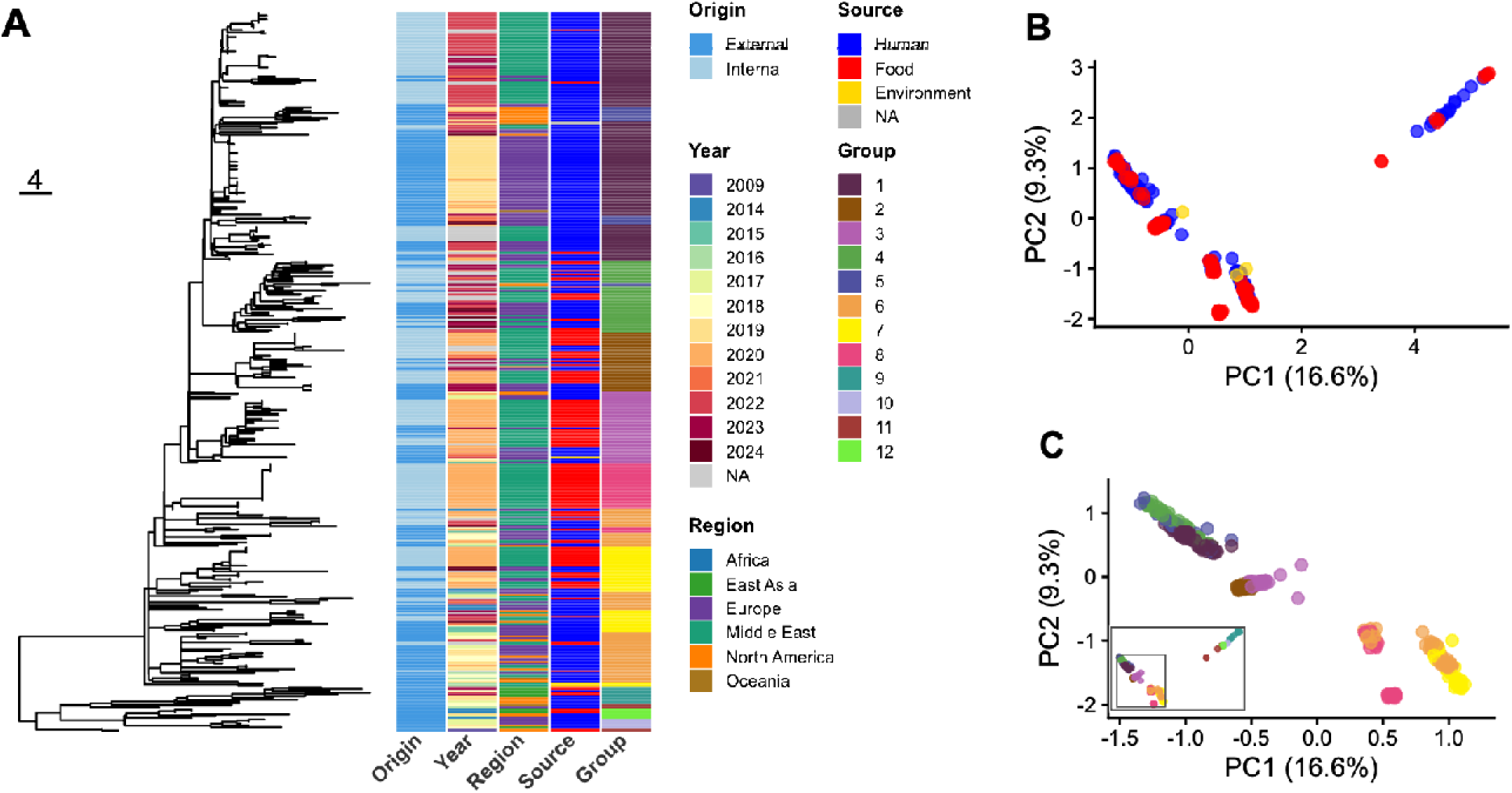
Population structure and phylogenetic relationships of 397 *Salmonella* Enteritidis isolates, comprising isolates from Saudi Arabia (internal collection) and globally sourced isolates that clustered with the Saudi collection (external collection). **(A)** Midpoint-rooted neighbor-joining phylogeny inferred from pairwise core-genome SNP distances across 1,218 variant sites; the scale bar denotes 4 SNPs. Metadata bars to the right of the tree indicate, from left to right, isolate origin (internal/external), year of isolation, geographic region, and BAPS group assignment. **(B)** Principal component analysis (PCA) of the same core-genome SNP genotypes, with isolates colored by source (human, food and environment). **(C)** The same ordination colored by Bayesian Analysis of Population Structure (BAPS) group. The main plot is restricted to BAPS groups 1–8, the inset shows the full ordination, with the grey box marking the magnified region.

### Recent sub-clones emerged from older ancestral lineages and exhibit increased antimicrobial resistance

We next examined the genetic, temporal, and antimicrobial resistance characteristics of each BAPS group (Figure 2). A neighbour-joining tree resolved the population into a series of recently emerged lineages (BAPS-1 to BAPS-8) connected to a smaller number of older and more genetically diverse lineages (BAPS-9 to BAPS-12). Saudi isolates were distributed across seven BAPS groups (BAPS-1 to BAPS-4 and BAPS-6 to BAPS-8), with no Saudi isolates assigned to BAPS-5 or BAPS-9 to BAPS-12 (Figure 2). Among these, BAPS-1 was the largest and most globally distributed group, comprising predominantly clinical isolates and linking Saudi clinical isolates with closely related genomes from Western Europe, North America and Oceania. In contrast, BAPS-2, BAPS-3 and BAPS-8 were significantly enriched for food-associated isolates (Fisher’s exact test, p-value < 0.05). The BAPS groups showed a clear temporal pattern. The predominantly non-Saudi groups (BAPS-10 to BAPS-12) consisted of older isolates, with a median sampling age of approximately 10 years, significantly older than the remainder of the population (Wilcoxon rank-sum test: BAPS-10 and BAPS-11, p-value < 0.01; BAPS-12, p-value < 0.001). In comparison, the recently emerged groups within the major genetic clade (BAPS-1 to BAPS-8) had a median isolate age of approximately 5 years. This temporal pattern was accompanied by marked differences in genomic diversity and antimicrobial resistance. Pairwise SNP distances revealed limited within-group diversity in several Saudi-associated groups, including BAPS-1, BAPS-3 and BAPS-8 (median, 1–9 SNPs), consistent with recent clonal expansion. In contrast, BAPS-4 to BAPS-7 and BAPS-9 exhibited significantly greater within-group diversity (median, 12–23 SNPs; p-value < 0.001), while BAPS-11 and BAPS-12 also showed higher diversity (median, 15–25 SNPs; p-value < 0.05), pointing to longer periods of diversification. Recently emerged groups also carried a significantly higher number of antimicrobial resistance determinants. BAPS-1 to BAPS-5 harboured more AMR genes than the remainder of the population (median, five genes; Wilcoxon rank-sum test, p-value < 0.001 for all comparisons). Together, these findings suggest that the dominant *S.* Enteritidis lineages circulating in Saudi Arabia emerged relatively recently from older ancestral populations and that this expansion has been accompanied by the accumulation of antimicrobial resistance determinants.

**Figure 2.**
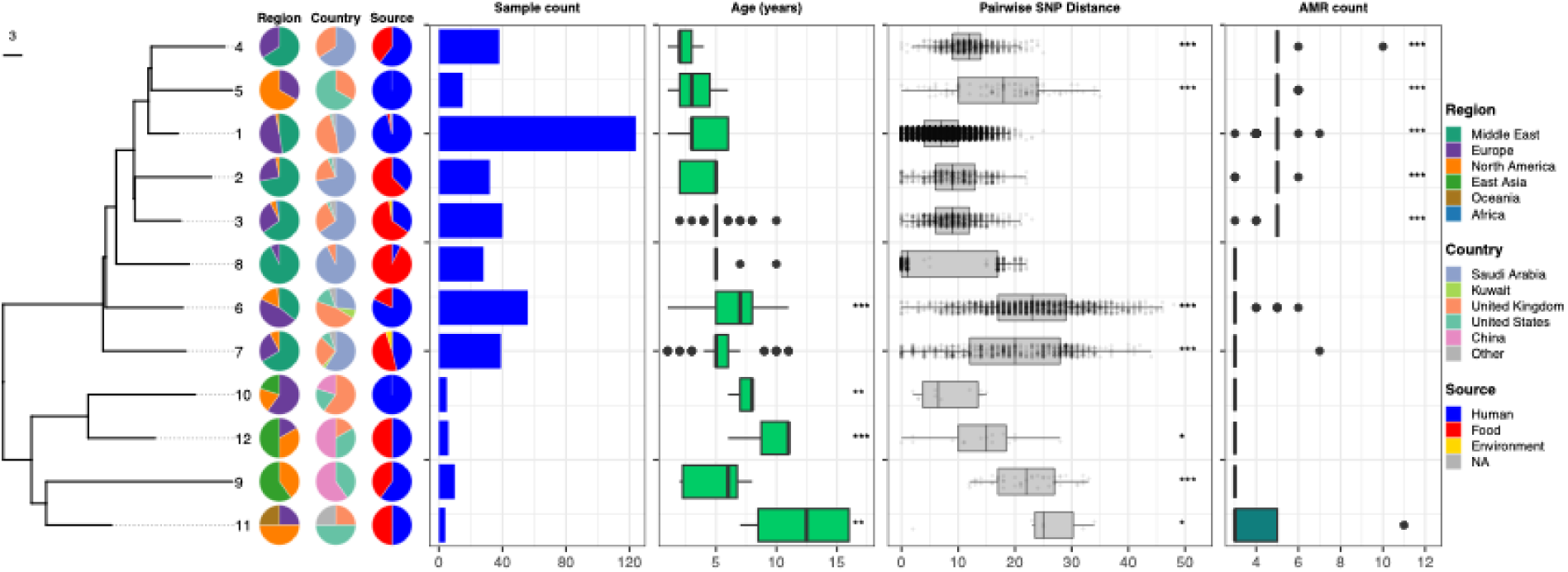
Population structure and genomic characteristics of the 12 *Salmonella* Enteritidis BAPS groups, comprising the Saudi Arabian collection and the related global isolates shown in **Figure 1**. The dendrogram (left) is a midpoint-rooted neighbor-joining (BIONJ) tree reconstructed from the mean pairwise core-genome SNP distances between genomes in different BAPS groups; branch lengths reflect genetic distance between groups, and the scale bar denotes 3 SNPs. The pie charts show the proportional composition of each group by geographic region, country and isolation source (human, food, environment). The blue bars show the number of genomes in each group. The boxplots for age show the distribution of the years of collection for the strains in each group. The boxplots for diversity correspond to the pairwise core-genome SNP distances between genomes within each group. The boxplots for the count of resistance genes show the distinct acquired antimicrobial resistance (AMR) genes identified by the genomic pipelines. Internal genomes are those sequenced as part of this study, while external genomes are from public databases (PathogenDetection). For the age, diversity and AMR panels, each BAPS group was compared with all remaining isolates by a one-sided Wilcoxon rank-sum test, with p-values adjusted across the 12 groups using the Benjamini–Hochberg procedure; the *, ** and *** signs denote adjusted significance levels of <0.05, <0.01 and <0.001, respectively.

### Repeated global introductions and subsequent local expansion of *S.* Enteritidis in the country

To further investigate the evolutionary dynamics of the population, we performed phylodynamic analyses on the five BAPS lineages (BAPS-1, BAPS-2, BAPS-3, BAPS-6, and BAPS-7) for which Bayesian inference achieved adequate convergence (Figure 3). The estimated ages of the most recent common ancestor (MRCA) indicated that these lineages emerged within the last 10– 15 years, with the two youngest lineages, BAPS-1 and BAPS-2, originating approximately 5–6 years ago (Figure 3A). Estimated nucleotide substitution rates were broadly consistent across lineages, ranging from approximately 4 × 10 to 1 × 10 substitutions per site per year (Figure 3A). These estimates are consistent with previous genomic studies reporting the recent emergence and clonal expansion of epidemic *S.* Enteritidis lineages (3, 5). Bayesian skyline analyses revealed rapid demographic expansion in all five groups, although two distinct demographic trajectories were observed (Figure 3B). BAPS-1 and BAPS-2 showed sustained increases in effective population size from their inferred origins to the present, whereas BAPS-3, BAPS-6, and BAPS-7 underwent an initial phase of rapid expansion followed by long-term endemic persistence (Figure 3B). To dissect the temporal and geographic relationships among isolates, we reconstructed time-scaled phylogenies for each group (Figure 3C). BAPS-1, BAPS-3, and BAPS-7 displayed phylogenetic structures consistent with one or more introduction events into the country followed by local clonal expansion. In these groups, the deepest reconstructed branches were predominantly associated with genomes from Northern Europe, particularly the United Kingdom, suggesting that these populations were established outside Saudi Arabia before becoming locally established (Figure 3C). Within BAPS-3, Saudi food-associated isolates were embedded within predominantly human-associated global clades. In contrast, BAPS-6 exhibited a more complex phylogeographic structure, comprising genomes from West Asia, i.e. Saudi Arabia, Kuwait, Lebanon, and other regions, consistent with repeated regional dissemination and population exchange rather than a single introduction event. BAPS-2 showed a distinct topology in which Saudi food-associated isolates served as ancestors to several UK genomes. Although incomplete global sampling precludes robust inference of transmission direction, this pattern is compatible with dissemination from a lineage circulating within the Saudi food reservoir into broader global populations, while alternative scenarios involving unsampled intermediary populations cannot be excluded. We also explored source-associated evolutionary dynamics within BAPS-2, BAPS-3, and BAPS-7 using Bayesian discrete-trait analyses to estimate transition rates between food and human sources. Despite uncertainty associated with the available sample sizes, the Bayesian discrete-trait analysis revealed a consistent pattern of source transitions across all three groups. All three groups yielded similar transition-rate estimates (BAPS-7: mean = 1.59 year ¹, 95% HPD = 0.02–4.99; BAPS-3: mean = 1.57 year ¹, 95% HPD = 0.04–4.89; BAPS-2: mean = 1.58 year ¹, 95% HPD = 0.02–5.02), although the broad HPD intervals indicate substantial uncertainty in the magnitude of these rates. Altogether, the phylogeographic analyses suggest that Saudi Arabia serves both as a recipient of globally circulating *S.* Enteritidis lineages and as a contributor to their subsequent regional and global dissemination.

**Figure 3.**
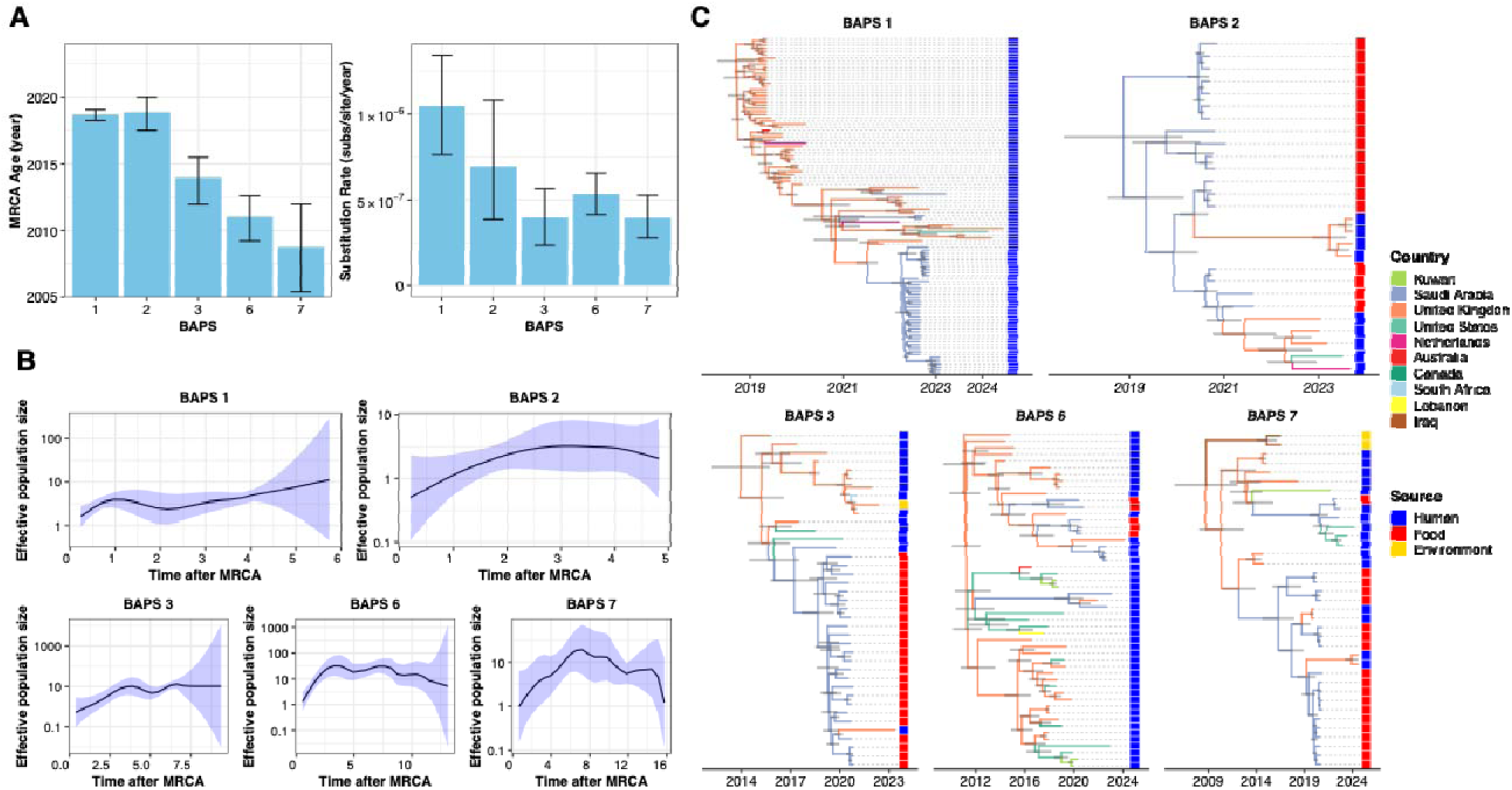
Phylodynamic reconstruction of the major *Salmonella* Enteritidis BAPS groups containing Saudi Arabian isolates. **(A)** Estimated most recent common ancestor (MRCA) age (left) and normalized nucleotide substitution rate (right) for each group, estimated by Bayesian analysis in BEAST. Error bars represent the 95% highest posterior density (HPD). **(B)** Skygrowth plot showing changes in effective population size over time. The solid line is the maximum a posteriori estimate and the shaded region indicates the 95% confidence interval. **(C)** Phylodynamic trees for the same clones, including genomes from the global collection. Internal branch colors denote the inferred country of origin, the tip strip denotes isolation source (human, food or environment), and horizontal grey bars represent the 95% HPD on internal node ages.

### Food reservoirs link human infection and global dissemination of S. Enteritidis

We reconstructed genomic connectivity networks using a 10-SNP threshold to identify clusters of closely related *S.* Enteritidis isolates and examine epidemiological links between reservoirs, focusing on BAPS groups containing both Saudi and global isolates. Across all groups, closely related isolates linked multiple geographic regions and source categories, with food-associated isolates frequently occupying central positions within the networks. BAPS-2, BAPS-3, BAPS-4, and BAPS-7 contained highly connected food-associated clusters, whereas human isolates were more often located at the network periphery (Figure 4). This pattern suggests that food reservoirs may represent hubs within the genomic connectivity network, although their apparent centrality may partly reflect greater sampling density. The persistence of *S.* Enteritidis within the connectivity networks was particularly evident in BAPS-4, where food isolates from central Saudi Arabia (Riyadh) appeared to serve as sources for infections diagnosed across geographically distant regions, including Qatif in the east, Najran in the south, and Asir in the southwest (Figure 4). Connectivity between food-associated and human-associated isolates was also observed across other groups, with closely related isolates frequently clustering together despite being collected from different sources, geographic regions, and years (Figure 4). Such connectivity patterns are consistent with the long-term persistence of *S.* Enteritidis lineages within food production and distribution systems and their recurrent association with human infections. The networks also revealed extensive dissemination within Saudi Arabia, with closely related isolates from BAPS-2, BAPS-3, BAPS-4, and BAPS-7 detected across multiple regions and spanning both human and food reservoirs (Figure 4). Together with the phylogenetic and phylodynamic analyses, these findings support repeated global introductions of *S.* Enteritidis into Saudi Arabia, followed by local expansion and dissemination across food and human reservoirs.

**Figure 4.**
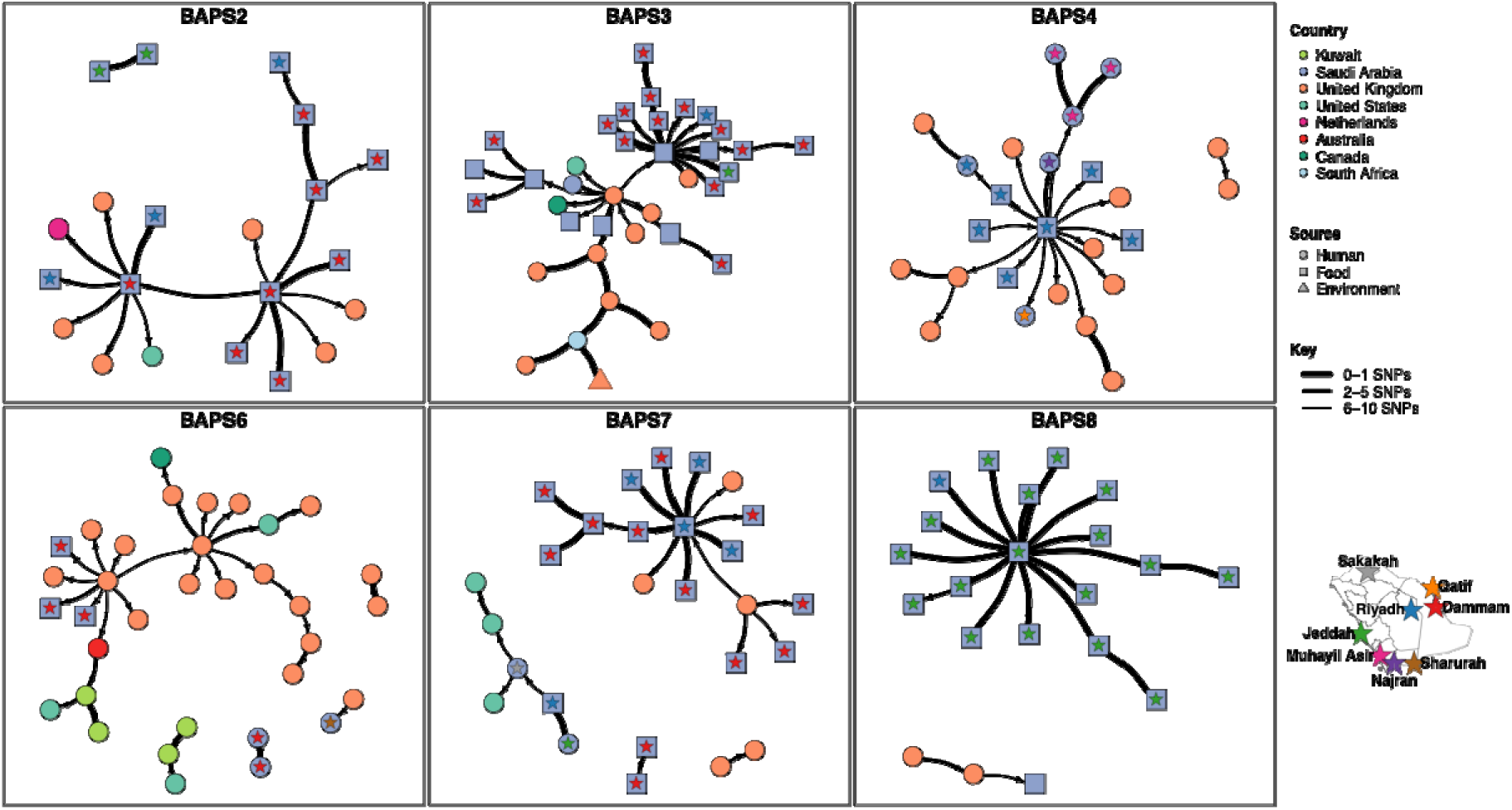
Connectivity and source-switching networks for the six BAPS clones containing both Saudi (internal) and global (external) isolates (BAPS 2, 3, 4, 6, 7 and 8). Each node represents one genome, colored by country of isolation, and edges link each genome to its most likely ancestor (inferred with seqTrack from pairwise core-genome SNP distances), with a distance cutoff for defining the networks. Node shape denotes source: circles, human; squares, food; triangles, environment. Arrows point from ancestor to descendant, and edge thickness reflects the pairwise SNP distance. A colored star on a node marks a Saudi isolate and encodes its city of origin, matching the inset map.

### Emergence of clones in the country was linked with increased antimicrobial resistance gene content

To investigate the genomic basis of the elevated antimicrobial resistance gene burden observed in recently emerged lineages (BAPS-1 to BAPS-5), we performed a comprehensive analysis of resistance determinants across all BAPS groups (Figure 5). Resistance determinants showed a heterogeneous distribution across the phylogeny, with evidence of multiple independent acquisition events rather than inheritance from a single resistant ancestor. While most lineages shared the conserved chromosomal efflux-associated genes *mdsA* and *mdsB*, substantial variation was observed in acquired resistance determinants. Moreover, fluoroquinolone resistance-associated mutations in *gyrA* showed a structured phylogenetic distribution. The quinolone resistance-determining region (QRDR) mutation *gyrA* D87Y was confined to the older lineages (BAPS-9 to BAPS-12), whereas *gyrA* S83Y was exclusively associated with the recently emerged clonal complex (BAPS-1 to BAPS-8) (Figure 5). In contrast, many acquired resistance genes showed lineage-specific or sporadic distributions. Aminoglycoside resistance genes, including *aph(3’’)-Ib* and *aph*(*6*)*-Id*, occurred in multiple clones, consistent with repeated gain and loss events. Similarly, several recent clones carried sporadically distributed β-lactamase genes, including the extended-spectrum β-lactamase (ESBL) determinants *bla*_CTX-M-8_ and *bla*_SHV-12_. The *bla*_CTX-M-8_ gene was identified in two isolates, including one food-associated isolate within BAPS-2 and one human-associated isolate within BAPS-4, whereas *bla*_SHV-12_ was detected in three human-associated isolates within BAPS-6. Although uncommon, the detection of clinically important ESBL determinants in both food and human isolates highlights the potential for shared resistance reservoirs across epidemiological sources. In contrast to ESBL genes, the narrow-spectrum β-lactamase *bla*_TEM-1_ was strongly associated with recently emerged clones and was present at high frequency in BAPS-1 to BAPS-5, where it occurred in nearly all isolates within each clone (Figure 5). A similar pattern was observed for *tetA*, which encodes a tetracycline efflux pump, with the gene enriched among recently expanding lineages but largely absent from the other clones. Together, the high prevalence of *bla*_TEM-1_, *tetA*, and other sporadically distributed resistance determinants suggests that the increased resistance of recently expanding *S.* Enteritidis lineages is linked to the acquisition of antimicrobial resistance determinants.

**Figure 5.**
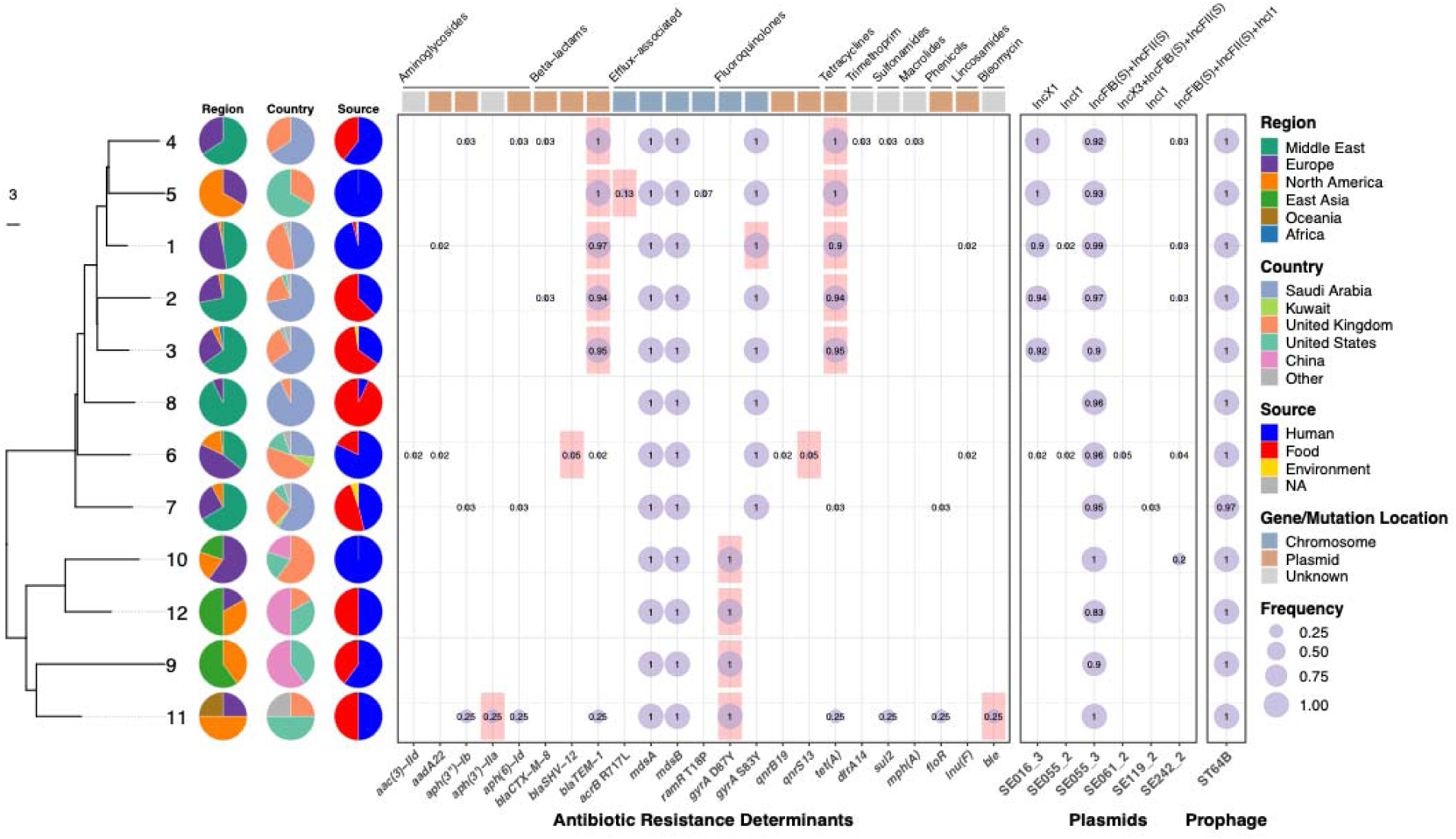
Distribution of antimicrobial resistance determinants, plasmids and the ST64B prophage across the *Salmonella* Enteritidis BAPS clones. Bubble sizes correspond to relative frequency, with the numbers indicating the relative frequency of each feature within each clone. The tree is the same as in Figure 2, and the stacked bars show the proportional composition of each clone by geographic region, country and isolation source (human, food, environment), as in Figure 2. Antimicrobial resistance determinants (genes and point mutations) are grouped by antimicrobial class (rotated labels along the top) and were identified using AMRFinderPlus. The coloured track above the resistance dots denotes the genomic context of each determinant: chromosome, plasmid, or unknown. The genomic context was determined based on the availability of full genome from long-read sequencing data. The shaded red squares indicate determinants significantly overrepresented in a clone compared with all other clones, as determined by a one-sided Fisher’s exact test with Benjamini–Hochberg correction and a significance level of adjusted P < 0.05. For the plasmid presence–absence patterns, short reads for each isolate were mapped against a representative plasmid backbone, using a cutoff of 90% coverage to determine presence of the plasmid; columns are labelled by Inc replicon type, and the genome IDs on the x-axis correspond to the isolate from which the reference plasmid genome was obtained. The ST64B prophage panel shows its prevalence in each clone on the same frequency scale.

### Plasmid-mediated dissemination underpins the emergence of antimicrobial resistance and virulence in *S.* Enteritidis clones

To determine the genomic context of antimicrobial resistance genes, we used hybrid long- and short-read sequencing to reconstruct plasmids from 25 representative isolates spanning all major BAPS groups and capturing the diversity of resistance gene and plasmid replicon profiles (see Methods). The analysis recovered 63 plasmids representing diverse replicon types and functional categories, including conjugative resistance plasmids and large resistance–virulence hybrid plasmids. Several plasmids carried acquired resistance determinants. Most notably, a 51.5-kb conjugative IncX1 plasmid carried the resistance genes *bla*_TEM-1_ and *tet(A)*, together with a putative virulence-associated YadA-like trimeric autotransporter adhesin and encoded a complete PilX/VirB type IV secretion system involved in conjugative transfer. This combination represents a dual risk, coupling resistance- and virulence-associated determinants with the capacity for horizontal dissemination (Figure S2A). This plasmid was highly prevalent in the recently emerged lineages BAPS-1 to BAPS-5 (90–100% of isolates in each group; Figure 5), explaining the enrichment of *bla*_TEM-1_ within these lineages. Its widespread distribution is consistent with the known mobility of IncX1 plasmids, which are self-transmissible, narrow-host-range replicons of *Enterobacteriaceae* previously reported in *S.* Enteritidis and recognized as key vehicles for the horizontal dissemination of resistance genes, supporting a potential role for this plasmid in the spread of resistance genes across expanding lineages in our collection (28, 31, 35). Besides the IncX1 plasmid, we identified the conserved serovar-associated virulence plasmid in isolates spanning all BAPS lineages (BAPS-1 to BAPS-12) (Figure 5). This 59.4-kb non-mobilizable IncFIB(S)/IncFII(S) plasmid carried the *spvRABCD* operon together with *pefA, pefC, pefD, rck*, and *mig-5* (Figure S2B). Its near-universal distribution is consistent with the conserved *S.* Enteritidis virulence plasmid carrying the *spv* operon, a key determinant of systemic infection (27). In addition to the commonly occurring plasmids, several plasmids were sporadically distributed across independent lineages. These included two large conjugative plasmids that combined the characteristic *S.* Enteritidis virulence backbone with acquired antimicrobial resistance determinants. A 103.8-kb IncFIB(S)/IncFII(S)/IncX3 plasmid carried the canonical virulence repertoire (*spvR, spvABC, pefA–D, pefI, rck*, and *mig-5*), together with the ESBL gene bla_SHV-12_ and the plasmid-mediated quinolone resistance gene *qnrS13* and was restricted to three clinical BAPS-6 isolates (Figures 5 and S3A). A second 149.7-kb conjugative IncFIB(S)/IncFII(S)/IncI1 plasmid carried a closely related virulence repertoire (*spvR, spvABCD, pefA–D, pefI, rck*, and *mig-5*) together with the ESBL gene *bla*_CTX-M-8_ and was identified in one food-associated Saudi BAPS-2 isolate and one UK clinical BAPS-4 isolate (Figures 5 and S3B). These plasmids represent hybrid virulence–resistance plasmids, consistent with fusion of the conserved IncFIB(S)/IncFII(S) *spv* virulence plasmid with resistance plasmids carrying acquired antimicrobial resistance genes, as reported before (36). Beyond IncX1, *bla*_TEM-1_ and *tet(A)* were also carried by two distinct IncI1 plasmid backbones. *bla*_TEM-1_ occurred on a 96.7-kb IncI1 plasmid together with *aadA22* and *lnu(F)*, identified in two clinical BAPS-1 isolates and one food-associated BAPS-6 isolate (Figure S3C). *tet(A)* was also carried on a 116-kb IncI1 plasmid within an MDR cassette comprising *tet(A), floR, aph*(*3*)*-Ib*, and *aph(6)-Id* in a clinical BAPS-7 isolate (Figure S3D). The plasmidome analysis revealed a highly dynamic plasmid landscape, with diverse resistance plasmids showing lineage-specific distributions and contributing to the spread of antimicrobial resistance alongside clonal expansion.

### Limited prophage diversity in the *S.* Enteritidis lineages

Having defined the plasmid landscape of the collection, we next characterized prophages across all major BAPS groups to obtain a broader view of the mobile accessory genome. Prophage screening for nine major classes of prophages in *Salmonella* (see Methods) identified a ST64B-like prophage (∼38% reference coverage) that was nearly ubiquitous across the population (Figure 5). Comparative analysis with the previously characterized ST64B reference (accession: AY055382.1) revealed conserved synteny across the major structural, integration, regulatory, and lysis modules (Figure S4), consistent with the lineage-associated distribution previously described for this prophage (37). ST64B is a lambdoid prophage originally characterized in *S.* Typhimurium, where its distribution has been linked to clonal lineage structure rather than ongoing horizontal exchange (37), while prophage content more broadly has been proposed as a useful marker of genomic diversity within *S.* enterica (38). The limited prophage diversity contrasts with the extensive plasmid variation, indicating that plasmids, rather than prophages, were the principal source of accessory genome diversity during the expansion of the *S.* Enteritidis population in the country.

## Discussion

In this study, we present the largest integrated food and human genomic analysis of *S.* Enteritidis ST11 from the Kingdom of Saudi Arabia and, to our knowledge, from West Asia. By integrating human and food-associated isolates collected over three years within a unified framework, we resolved the population structure, reconstructed patterns of global dissemination and cross-reservoir transmission and characterized the mobile genetic elements underlying the spread of antimicrobial resistance. Overall, our findings indicate that the *S.* Enteritidis ST11 population in Saudi Arabia forms part of a globally circulating epidemiological cluster that has diversified into several expanding subclones with elevated resistance and virulence. These lineages appear to have been shaped by repeated global introductions, establishment within poultry-associated reservoirs, and the acquisition of resistance- and virulence-associated plasmids. These findings highlight the value of integrated food and clinical surveillance while underscoring mobile genetic elements as key targets for future genomic surveillance of this lineage.

Our results showed that the *S.* Enteritidis population in Saudi Arabia was embedded within multiple globally distributed epidemic lineages spanning Europe, North America, Oceania, the Middle East and Africa, consistent with repeated global introductions followed by local expansion. This pattern agrees with previous genomic studies in other regions demonstrating repeated introductions and subsequent local establishment of *S.* Enteritidis clones (4), although nationwide genomic studies have also shown that geography remains an important determinant of population structure (18, 20). Repeated introductions likely reflect Saudi Arabia’s position as a major hub for both international food trade and human mobility. Global trade in poultry breeding stocks has been widely implicated in the worldwide dissemination of epidemic *S.* Enteritidis lineages (3), while the high levels of international human mobility associated with migration and mass gatherings, particularly the Hajj and Umrah pilgrimages, may provide additional opportunities for the introduction and dissemination of globally circulating strains (34, 39). The relative importance of these introduction pathways warrants further investigation using integrated genomic, epidemiological, and food-chain surveillance. Following their introduction, however, our results suggest that these lineages are maintained within the country through continued circulation between poultry-associated reservoirs and humans, rather than forming independent human- and food-associated populations.

Our study reports the lineage-specific expansion of a conjugative IncX1 plasmid carrying *bla*_TEM-1_ within the recently emerged clones. Plasmid-mediated dissemination of resistance genes is well established across *Enterobacterales* (28, 35), and our hybrid assembly approach provides a high-resolution reconstruction of the *S.* Enteritidis ST11 plasmidome. The near-fixation of IncX1 within these recently expanded Saudi-associated lineages fits within a broader pattern of IncX-mediated resistance evolution observed globally. This finding supports the growing evidence that IncX1-associated resistance evolution is geographically structured rather than uniformly distributed across the global ST11 population. Comparative analyses of human isolates from China, the United States, Europe and Africa found IncX1 almost exclusively among multidrug-resistant Chinese isolates, with alternative plasmid families predominating elsewhere (12), while a large Spanish genomic study similarly identified IncX1 within a distinct resistance-enriched clade that remained largely separate from a susceptible human-associated lineage (20). Poultry-associated genomic surveillance from China also identified IncX1 as a major carrier of acquired resistance genes, including *bla*_CTX-M-14_*, fosA3, qnrS1*, and *tet(A)* (40, 41), while surveillance in

Singapore identified a shared IncFIB(S)-IncFII(S)-IncX1 plasmid profile in both poultry and human ST11 isolates, directly linking farm and clinical populations (42). A longitudinal study of Chinese poultry production demonstrated that the IncX1 plasmid backbone was disseminated throughout the poultry production chain and that antimicrobial resistance differences among closely related bacterial lineages were largely attributable to plasmid acquisition (25). Taken together, these studies indicate that IncX1 can disseminate within established *S.* Enteritidis populations, particularly through poultry-associated reservoirs, and subsequently contribute to the emergence of resistant sublineages. Consistent with this, our findings suggest that IncX1 was acquired relatively recently by an already globally disseminated *S.* Enteritidis lineage and contributed to the elevated resistance of the recently emerged Saudi-associated clones, rather than driving the initial emergence or global dissemination of the lineage itself. We also observed that the *gyrA* S83Y mutation was confined to the recently emerged IncX1-positive clonal complex, whereas older IncX1-negative lineages predominantly carried the *gyrA* D87Y mutation. A recent study provides a possible framework for interpreting this pattern, demonstrating that the distribution of *gyrA* mutations in *S.* Enteritidis is associated with the carriage of IncX1 and the ESSI-2 bacteriophage, and that these relationships may be further shaped by the poultry litter microbial community (43). Although the direction of these associations differed from that observed in our population, these findings support the broader hypothesis that interactions between mobile genetic elements, chromosomal resistance mutations, and the poultry-associated ecological environment may influence the evolutionary trajectories of fluoroquinolone resistance in *S.* Enteritidis.

The recurrence of closely related plasmid backbones across multiple phylogenetic lineages and across both food and human reservoirs further indicates that horizontal gene transfer has played a major role in shaping the evolution of this population. Rather than representing multiple independent acquisition events, these shared plasmids are more consistent with circulation within a common mobile gene pool, highlighting the well-known role of conjugative plasmids in disseminating resistance across *Enterobacterales* (25, 28, 30–32, 35, 36). The identification of hybrid virulence-resistance plasmids that co-located resistance determinants and canonical virulence loci on the same backbone is particularly concerning. Such genetic architectures may facilitate the co-selection and co-dissemination of resistance and virulence under antimicrobial pressure. Their occurrence across multiple phylogenetic lineages and ecological reservoirs, together with their close similarity to hybrid plasmids reported from other *S.* Enteritidis and *Enterobacterales* populations (31, 36), suggests that these elements form part of a broader circulating mobile genetic pool rather than representing isolated evolutionary events. These findings suggest that surveillance should target mobile genetic elements alongside bacterial clones.

Despite the depth of our collection, we note several limitations. First, our collection included a limited number of environmental samples and no animal samples, both of which are key components of a One Health surveillance programme. Second, we analysed predominantly MDR strains from hospitals. Although this allowed us to identify the role of distinct antimicrobial resistance genes in the emergence of new clones, we could not examine the other evolutionary forces that may have contributed to their emergence, including host and ecological adaptation, virulence-associated selection, and demographic processes, e.g. founder effects and genetic drift. The inclusion of a broader collection that includes susceptible strains would help to establish whether the overall population structure of *S.* Enteritidis remains consistent with our results. Finally, our study spanned only three years, limiting our ability to investigate the longer-term population dynamics of *S.* Enteritidis. This relatively short sampling period may also explain the weak temporal signal and the broad credible intervals for key phylodynamic parameters, including transition rates between human and food reservoirs. Continued longitudinal genomic surveillance will be needed to improve these estimates and to better characterize the long-term evolution and spread of the expanding lineage. This will improve the precision of phylodynamic inferences, enable more robust reconstruction of transmission pathways, and provide stronger evidence to guide public health interventions.

## Methods

### Sample collection and isolation

The overall workflow of the study is summarized in Figure S1. Over the course of the study, the Microbiology Reference Laboratory at the Saudi Food and Drug Authority (SFDA) received Salmonella isolates from processed and unprocessed chicken products through national food surveillance and monitoring programmes conducted according to World Health Organization sampling guidelines. Among these, 123 isolates were identified as *Salmonella enterica* serovar Enteritidis (*S.* Enteritidis) and included in this study. Food isolates were generated by ISO/IEC 17025-accredited laboratories, including SFDA control laboratories in Riyadh, Jeddah, and Dammam, as well as laboratories of the Ministry of Environment, Water and Agriculture (MEWA), Saudi Arabia. All laboratories followed the ISO 6579-1:2017 standard for the detection, isolation, and serotyping of *Salmonella* from food samples. Positive isolates were submitted to the SFDA Reference Laboratory of Microbiology for confirmatory identification by MALDI-TOF mass spectrometry, serotyping according to the Kauffmann–White scheme using commercial O antisera (Remel, UK), and molecular confirmation using the SureFast *Salmonella* PLUS real-time PCR assay (R-Biopharm, Germany). Confirmed isolates were archived in the SFDA biobank at −80°C until further analysis.

Clinical isolates were collected and characterized using the same laboratory framework as food isolates. A total of 97 *Salmonella enterica* serovar Enteritidis (*S.* Enteritidis) isolates were obtained from patients with laboratory-confirmed salmonellosis diagnosed through routine clinical practice at five Ministry of Health (MOH) diagnostic laboratories across Saudi Arabia between 2022 and 2023. Most isolates were recovered from stool specimens (n=72), with the remainder obtained from other clinical samples, including blood cultures. Stool and rectal swab specimens were cultured on MacConkey, Hektoen Enteric, and Xylose Lysine Deoxycholate (XLD) agars and incubated at 37 °C for 18–24 h. Blood cultures were processed using the BacT/ALERT® automated system (bioMérieux), and positive cultures were subcultured under standard microbiological conditions. Presumptive *Salmonella* isolates were identified using the VITEK® 2 system (bioMérieux, Marcy-l’Étoile, France) before being transported under controlled conditions to the SFDA Reference Laboratory of Microbiology (RLM). At the RLM, all isolates underwent confirmatory identification and serotyping according to the Kauffmann– White scheme to verify *S.* Enteritidis. Following confirmation, both food and clinical isolates were archived in the RLM biobank at −80 °C until whole-genome sequencing and downstream analyses. Antimicrobial susceptibility testing of the isolates was performed using the broth microdilution method with the Sensititre EUVSEC3 panel (TREK Diagnostic Systems, Thermo Fisher Scientific), following the manufacturer’s instructions for inoculation and incubation.

### DNA extraction and whole genome sequencing with short-read sequencing

Genomic DNA was isolated using the DNeasy Blood & Tissue Kit (Qiagen, CA, USA) following the manufacturer’s recommended procedure. DNA quality was evaluated using the QIAxpert system (Qiagen), with an A260/A280 ratio of ≥1.8 considered acceptable, while DNA concentration was determined using a Qubit fluorometer (Invitrogen, CA, USA). Sequencing libraries were generated using Illumina DNA library preparation reagents and FX Index Kits (Illumina, San Diego, CA, USA). After index PCR, library purification was performed using 45 μL of Agencourt AMPure XP magnetic beads (Beckman Coulter, Brea, CA, USA) at a 3:2 sample-to-bead ratio. Purified libraries were quantified by Qubit, normalized to comparable concentrations, and pooled for sequencing. The pooled libraries were subsequently sequenced on an Illumina NovaSeq 6000 platform using 250-bp paired-end reads and a 5% PhiX spike-in control, in accordance with the manufacturer’s guidelines.

### Short-read sequencing data analysis

After sequencing, raw FASTQ reads were quality-controlled using fastQC (v.0.11.3) (http://www.bioinformatics.babraham.ac.uk/projects/fastqc/) and Trimmomatic (v.0.36) (44). Filtered short reads were then processed through the downstream pipeline. *De novo* assemblies were generated using Unicycler (v0.5.1) with default parameters (45) (https://github.com/rrwick/Unicycler), followed by genome annotation using Bakta (v1.12.0) (46). Assembly quality was assessed using QUAST (v5.2.0) (47) and sequence type was assigned using MLST (v2.23.0) (https://github.com/tseemann/mlst). Antimicrobial resistance (AMR) determinants were identified using AMRFinderPlus (v3.12.8) (48). Plasmid-associated sequences were identified using ABRicate (v1.0.1) (https://github.com/tseemann/abricate) and PlasmidHunter (49) against the PlasmidFinder database (50). For ABRicate-based screening, minimum thresholds of 90% sequence identity and 90% coverage were applied. Resistance mutations were further confirmed using staramr v0.7.1 (https://github.com/phac-nml/staramr) (51). The tool was also used to predict drug-resistant phenotypes.

### Contextualization of the samples

To contextualize the Saudi genome collection, we queried the NCBI Pathogen Detection database on 15 March 2026 for all publicly available *Salmonella enterica* serovar Enteritidis ST11 genomes. The Saudi isolates were predominantly assigned to a single NCBI Pathogen Detection SNP cluster (PDS000026888.159). At the time of analysis, Salmonella enterica clusters in the NCBI Pathogen Detection database were generated using the whole-genome multilocus sequence typing (wgMLST) clustering pipeline, in which genomes are grouped by single-linkage clustering using a maximum of 25 wgMLST allele differences between linked genomes. Within each cluster, reference-based SNP calling is subsequently performed to infer phylogenetic relationships among closely related isolates (www.ncbi.nlm.nih.gov/pathogens/docs/data_processing). We retrieved all publicly available external genomes within this cluster that had complete metadata, including isolation source, country, and collection date, and combined them with the Saudi genomes for comparative analyses, yielding a final comparative dataset for downstream analyses. Core-genome SNPs were identified by aligning reads to the *S.* Enteritidis PT4 strain P125109 reference genome (AM933172.1) using Snippy (v4.6.0) (https://github.com/tseemann/snippy). Pairwise SNP distances were calculated from the core-genome SNP alignment, and a neighbour-joining tree was constructed using the R package ape (v5.8.1) (52). The resulting phylogeny and the metadata were visualised using the R package ggtree (v4.0.5) (53). Population structure of *S.* Enteritidis ST11 was inferred from pairwise SNP distances using RhierBAPS (54) with a single clustering level. We next constructed a minimum spanning tree of 12 identified BAPS lineages (BAPS-1 to BAPS-12) by calculating the mean pairwise SNP distances between genomes from different BAPS lineages. The generated average distance matrix was then used to construct a neighbour-joining tree using the R package ape (v5.8.1). Details of all genomes are provided in Supplementary Table S1. The genomic accession numbers and associated metadata are also available through the Saudi Pathogen Genome Atlas (https://saudipathogenatlas.kaust.edu.sa/).

### Phylodynamic analysis

To examine the global epidemiological structure of *S.* Enteritidis ST11, phylodynamic analyses were conducted on BAPS lineages containing Saudi genomes (BAPS-1 to BAPS-8, except BAPS-5). For each BAPS lineage, short-reads from all strains were mapped to the local reference genome using Snippy (v4.6.0) (https://github.com/tseemann/snippy), producing core-genome SNP alignments. Each local reference was assembled by concatenating contigs from the genome with the highest N50 in the BAPS group. Following read mapping, SNP sites were extracted from the core-genome SNP alignments, and hypervariable regions were removed using Gubbins (v3.3.1) (55) with five iterations. The resulting recombination-filtered SNP alignments were then used as input for Bayesian Evolutionary Analysis Sampling Trees (BEAST) v1.10.4 (56) to perform phylogeographic diffusion analysis. In doing so, the geographical location, defined as the country of isolation, was assigned as a discrete trait for each genome. A constant population size model was applied, with a uniform distribution specified for the clock rate. For the discrete-trait analysis, a symmetric substitution model with a uniform distribution was used. The Markov chain Monte Carlo (MCMC) chain was run for 10^9^ iterations. Convergence, defined as an effective sample size (ESS) >150 for all key parameters, was achieved for BAPS-1, BAPS-2, BAPS-3, BAPS-6, and BAPS-7, but not for the remaining BAPS lineages. TreeAnnotator, part of the BEAST package, was used to generate dated phylogenetic trees, which were visualized using the R package ggtree (v4.0.5). To estimate the rate of transfer of food-human sources, we also repeated the above analysis, but source of isolation (i.e. food or human) was assigned as a discrete trait for each genome. To assess changes in population size over time, a non-parametric growth model was applied to estimate effective population size along the time-scaled phylogenies of the five BAPS groups for which dated trees could be obtained. This analysis was performed using the skygrowth.map function, and the results were visualized using the plot function in the R package skygrowth v0.3.1 (57).

### Genomic connectivity analysis

To infer putative recent genomic connectivity between human and food reservoirs, maximum-likelihood transmission network analyses integrating pairwise SNP distances and isolate sampling dates were performed for the selected BAPS clusters (BAPS-2, BAPS-3, BAPS-4, BAPS-6, BAPS-7, and BAPS-8) using the R package adegenet (58). These groups contained both human and food-associated genomes, with at least one genome originating from the Saudi collection. For each BAPS group, recombination-filtered SNP alignments generated above were used to calculate pairwise genetic distances using the dist.dna function from the R package ape with default settings. These distance matrices and isolation dates were then used as input to the seqTrack function in the adegenet package in R. Genomic connectivity networks were defined using a 10-SNP threshold (59), representing broadly related *Salmonella* genomes, and were used to identify genomic clusters for surveillance purposes. The inferred networks were visualised using the R package igraph (v2.3.2) (60).

### Plasmidome analysis

To characterize the plasmidome across the genome collection, third-generation long-read sequencing was performed on representative isolates selected to capture the phylogenetic diversity of the collection. Representative isolates were selected from each BAPS group containing at least one Saudi isolate. We selected 25 *S.* Enteritidis ST11 strains based on genome quality and the presence of at least one AMR gene and one plasmid replicon. Genomic DNA concentration and purity were assessed using a Qubit HS dsDNA kit and spectrophotometry at 260/280 nm. Multiplexed libraries were prepared using 96-plex Rapid Barcoding Kits and sequenced on MinION flow cells (Oxford Nanopore Technologies) according to the manufacturer’s instructions for a 48-hour sequencing run. Genomes were sequenced using Oxford Nanopore Technologies (ONT) long-read sequencing and assembled using a hybrid approach that combined long and short read sequencing data using Unicycler (v0.4.7) (45, 61) with default parameters. Hybrid assemblies were assessed and visualized for quality using QUAST (v5.2.0) (47) with default parameters and Bandage (v0.9.0) (62). Plasmid fragments were extracted using Bandage (v0.9.0), and plasmid incompatibility groups were assigned by BLAST of the extracted plasmid sequences against the PlasmidFinder database. To characterize the plasmid content of the population, all plasmid fragments containing a known plasmid replicon were extracted and grouped by incompatibility (Inc) type. A representative plasmid from each incompatibility group was selected based on size and annotated and visualized using Bakta (v1.12.0) and Proksee (v1.2.1) (63), respectively. The built-in tools in Proksee were used to identify origins of replication, AMR and virulence genes. To examine the phylogenetic distribution of these plasmids across the entire *S.* Enteritidis ST11 population, each representative plasmid was queried against the assembled genome of every strain using BLASTn (64). Plasmid presence was defined by a minimum sequence coverage threshold of 90%.

### Prophage analysis

To characterize the prophage repertoire of the population, nine previously described *Salmonella* prophage reference sequences (Fels-1 (NC_010391.1), Fels-2 (NC_010463.1), Gifsy-1 (NC_010392.1), Gifsy-2 (NC_010393.1), P22 (NC_002371.2), RE-2010 (NC_019488.1), SopE (AY319521.1), ST64B (AY055382.1) and ST64T (AY052766.1)) were screened against the assembled genomes of all isolates using BLASTn (≥70% nucleotide identity and ≥30% query coverage). Of the nine prophages, only ST64B exhibited variable presence across the entire collection and was therefore selected for further analysis. A representative ST64B prophage was annotated using Bakta (v1.12.0) and prophage regions were identified and visualized with PHASTEST (v3.0) (65) in Proksee (v1.2.1) to define prophage boundaries and annotate gene content.

### Statistical significance tests

One-sided Fisher’s exact tests were used for AMR determinant enrichment, and one-sided Wilcoxon rank-sum tests for differences in age, SNP diversity and AMR gene count between BAPS groups. P values were adjusted by the Benjamini–Hochberg method in R (v4.4.2). Associations between isolate source (clinical or food) and antimicrobial resistance status were assessed by classifying isolates as MDR, non-MDR resistant, or susceptible. MDR was defined as predicted resistance to at least one antimicrobial agent in three or more antimicrobial classes, whereas non-MDR isolates exhibited predicted resistance but did not meet the MDR definition. Isolates with no predicted resistance were classified as susceptible. The overall association between isolate source and resistance status was assessed using Pearson’s chi-squared test and Fisher’s exact test. Statistical analyses were performed in R. For the one-sided tests described above, an adjusted p < 0.05 was considered statistically significant; the association between isolate source and resistance status was assessed using two-sided tests at p < 0.05.

## Data Availability

The short- and long-read sequencing data were submitted to the European Nucleotide Archive (ENA) under study number PRJEB79567. Metadata, profiles of antimicrobial resistance genes/mutations, virulence factor genes, and plasmid replicons, as well as the accession data for external (global) genomes, are provided in Supplementary Table S1. All other intermediate files and relevant tables are available on the GitHub directory for the project: https://github.com/gzhoubioinf/SalmonellaEnteritidis_SFDA.git.

## Acknowledgements

G.Z., J.H., S.I., N.H., O.F., and D.M. were supported by baseline funding from King Abdullah University of Science and Technology (KAUST) (BAS/1/1108-01-01) and by the KAUST Center of Excellence for Smart Health (FCC/1/5932-01-03). M.B. was supported by a UKRI Future Leaders Fellowship (MR/V027204/1). The authors gratefully acknowledge the Saudi Food and Drug Authority (SFDA) for providing access to isolates, metadata, and laboratory resources that made this study possible.

## Author Contributions

F.A., G.Z., and J.H. contributed equally to this work and share first authorship. F.A. conducted the experimental and computational analyses. G.Z. and J.H. contributed to data analysis, bioinformatics, and interpretation of the results. N.H., A.A., N.M.A., M.S.A., K.O.A., S.A.S., M.A.A., A.A.A., E.A.A., A.A.T., H.H.A., S.I.A., A.A.A., A.L.A., M.S.R., M.B. and S.A. contributed samples, laboratory resources, epidemiological data, and technical expertise. S.I., and O.F. contributed to data analysis. L.M., S.F., and D.M. contributed to the conceptualisation of the study. L.M., S.F., and D.M. supervised the study. F.A., G.Z., and D.M. wrote the manuscript with input from all authors. All authors reviewed, edited, and approved the final manuscript.

## Ethical approvals

This study was approved by the Institutional Biosafety and Bioethics Committee (IBEC) of King Abdullah University of Science and Technology (Approval No. 22IBEC046) and the Institutional Review Board (IRB) of the General Administration of Research and Studies, Ministry of Health (MOH), Saudi Arabia (Approval No. 23-23 M).

## References

1. Majowicz SE, Musto J, Scallan E, Angulo FJ, Kirk M, O’Brien SJ, et al. The global burden of nontyphoidal Salmonella gastroenteritis. Clin Infect Dis. 2010;50(6):882–9.

2. Kirk MD, Pires SM, Black RE, Caipo M, Crump JA, Devleesschauwer B, et al. World Health Organization Estimates of the Global and Regional Disease Burden of 22 Foodborne Bacterial, Protozoal, and Viral Diseases, 2010: A Data Synthesis. PLoS Med. 2015;12(12):e1001921.

3. Li S, He Y, Mann DA, Deng X. Global spread of Salmonella Enteritidis via centralized sourcing and international trade of poultry breeding stocks. Nat Commun. 2021;12(1):5109.

4. Luo L, Payne M, Kaur S, Hu D, Cheney L, Octavia S, et al. Elucidation of global and national genomic epidemiology of Salmonella enterica serovar Enteritidis through multilevel genome typing. Microb Genom. 2021;7(7):000605.

5. Feasey NA, Hadfield J, Keddy KH, Dallman TJ, Jacobs J, Deng X, et al. Distinct Salmonella Enteritidis lineages associated with enterocolitis in high-income settings and invasive disease in low-income settings. Nat Genet. 2016;48(10):1211–7.

6. Alghoribi MF, Doumith M, Alrodayyan M, Al Zayer M, Köster WL, Muhanna A, et al. S. Enteritidis and S. Typhimurium Harboring SPI-1 and SPI-2 Are the Predominant Serotypes Associated With Human Salmonellosis in Saudi Arabia. Front Cell Infect Microbiol. 2019;9:187.

7. European Food Safety Authority, European Centre for Disease Prevention and Control. The European Union One Health 2021 Zoonoses Report. EFSA J. 2022;20(12):e07666.

8. Sher AA, Mustafa BE, Grady SC, Gardiner JC, Saeed AM. Outbreaks of foodborne Salmonella enteritidis in the United States between 1990 and 2015: An analysis of epidemiological and spatial-temporal trends. Int J Infect Dis. 2021;105:54–61.

9. Al-Rifai RH, Chaabna K, Denagamage T, Alali WQ. Prevalence of enteric non-typhoidal Salmonella in humans in the Middle East and North Africa: A systematic review and meta-analysis. Zoonoses Public Health. 2019;66(7):701–28.

10. Rodrigue DC, Tauxe RV, Rowe B. International increase in Salmonella enteritidis: a new pandemic? Epidemiol Infect. 1990;105(1):21–7.

11. Achtman M, Wain J, Weill FX, Nair S, Zhou Z, Sangal V, et al. Multilocus sequence typing as a replacement for serotyping in Salmonella enterica. PLoS Pathog. 2012;8(6):e1002776.

12. Cao G, Zhao S, Kuang D, Hsu CH, Yin L, Luo Y, et al. Geography shapes the genomics and antimicrobial resistance of Salmonella enterica Serovar Enteritidis isolated from humans. Sci Rep. 2023;13(1):1331.

13. Zhou H, Jia C, Shen P, Huang C, Teng L, Wu B, et al. Genomic census of invasive nontyphoidal Salmonella infections reveals global and local human-to-human transmission. Nat Med. 2025;31(7):2325–34.

14. Abdelhamid AG, Yousef AE. Egg-associated Salmonella enterica serovar Enteritidis: comparative genomics unveils phylogenetic links, virulence potential, and antimicrobial resistance traits. Front Microbiol. 2023;14:1278821.

15. Di Marcantonio L, Janowicz A, Zilli K, Romantini R, Bilei S, Paganico D, et al. Genomic Comparison of Salmonella Enteritidis Strains Isolated from Laying Hens and Humans in the Abruzzi Region during 2018. Pathogens. 2020;9(5):349.

16. Hyeon JY, Li S, Mann DA, Zhang S, Kim KJ, Lee DH, et al. Whole-Genome Sequencing Analysis of Salmonella Enterica Serotype Enteritidis Isolated from Poultry Sources in South Korea, 2010-2017. Pathogens. 2021;10(1):45.

17. Leao C, Silveira L, Usie A, Giao J, Clemente L, Themudo P, et al. Genetic Diversity of Salmonella enterica subsp. enterica Serovar Enteritidis from Human and Non-Human Sources in Portugal. Pathogens. 2024;13(2):112.

18. Toro M, Retamal P, Ayers S, Barreto M, Allard M, Brown EW, et al. Whole-Genome Sequencing Analysis of Salmonella enterica Serovar Enteritidis Isolates in Chile Provides Insights into Possible Transmission between Gulls, Poultry, and Humans. Appl Environ Microbiol. 2016;82(20):6223–32.

19. Usein CR, Oprea M, Ciontea AS, Dinu S, Cristea D, Zota LC, et al. A Snapshot of the Genetic Diversity of Salmonella Enteritidis Population Involved in Human Infections in Romania Taken in the European Epidemiological Context. Pathogens. 2021;10(11):1490.

20. Samper-Cativiela C, Torre-Fuentes L, Diéguez-Roda B, Maex M, Ugarte-Ruiz M, Carrizo P, et al. Molecular epidemiology of Salmonella Enteritidis in humans and animals in Spain. Antimicrob Agents Chemother. 2025;69(4):e0073824.

21. Yan W, Xu D, Chen L, Wu X. Antimicrobial resistance and genome characteristics of Salmonella enteritidis from Huzhou, China. PLoS One. 2024;19(6):e0304621.

22. Aldrich C, Hartman H, Feasey N, Chattaway MA, Dekker D, Al-Emran HM, et al. Emergence of phylogenetically diverse and fluoroquinolone resistant Salmonella Enteritidis as a cause of invasive nontyphoidal Salmonella disease in Ghana. PLoS Negl Trop Dis. 2019;13(6):e0007485.

23. Castro-Vargas RE, Herrera-Sánchez MP, Rodríguez-Hernández R, Rondón-Barragán IS. Antibiotic resistance in Salmonella spp. isolated from poultry: A global overview. Vet World. 2020;13(10):2070–84.

24. Wang Y, Xu X, Jia S, Qu M, Pei Y, Qiu S, et al. A global atlas and drivers of antimicrobial resistance in Salmonella during 1900-2023. Nat Commun. 2025;16(1):4611.

25. Lei CW, Zhang Y, Kang ZZ, Kong LH, Tang YZ, Zhang AY, et al. Vertical transmission of Salmonella Enteritidis with heterogeneous antimicrobial resistance from breeding chickens to commercial chickens in China. Vet Microbiol. 2020;240:108538.

26. D’Alessandro B, Pérez Escanda V, Balestrazzi L, Grattarola F, Iriarte A, Pickard D, et al. Comparative genomics of Salmonella enterica serovar Enteritidis ST-11 isolated in Uruguay reveals lineages associated with particular epidemiological traits. Sci Rep. 2020;10(1):3638.

27. Silva C, Puente JL, Calva E. Salmonella virulence plasmid: pathogenesis and ecology. Pathog Dis. 2017.

28. Zhang XZ, Lei CW, Zeng JX, Chen YP, Kang ZZ, Wang YL, et al. An IncX1 plasmid isolated from Salmonella enterica subsp. enterica serovar Pullorum carrying bla(TEM-1B), sul2, arsenic resistant operons. Plasmid. 2018;100:14–21.

29. Graham RMA, Hiley L, Rathnayake IU, Jennison AV. Comparative genomics identifies distinct lineages of S. Enteritidis from Queensland, Australia. PLoS One. 2018;13(1):e0191042.

30. Yang S, Fan J, Yu L, He J, Zhang L, Yu Y, et al. Dissemination of Ceftriaxone-Resistant Salmonella Enteritidis Harboring Plasmids Encoding bla(CTX-M-55) or bla(CTX-M-14) Gene in China. Antibiotics (Basel). 2024;13(5).

31. Liu YY, He DD, Zhang MK, Pan YS, Wu H, Yuan L, et al. The Formation of Two Hybrid Plasmids Mediated by IS26 and Tn6952 in Salmonella enterica Serotype Enteritidis. Front Microbiol. 2021;12:676574.

32. Huang J, Alzahrani KO, Zhou G, Alsalman SA, Alsufyani AT, Alotaibi NM, et al. Genomic survey of multidrug resistant Salmonella enterica serovar Minnesota clones in chicken products. NPJ Antimicrob Resist. 2025;3(1):10.

33. General Authority for Statistics (GASTAT). Food Security Statistics for 2023. Riyadh, Saudi Arabia: General Authority for Statistics, Kingdom of Saudi Arabia; 2024.

34. Aldossari M, Aljoudi A, Celentano D. Health issues in the Hajj pilgrimage: a literature review. East Mediterr Health J. 2019;25(10):744–53.

35. Zalewska M, Blazejewska A, Gawor J, Adamska D, Goryca K, Szelag M, et al. The IncC and IncX1 resistance plasmids present in multi-drug resistant Escherichia coli strains isolated from poultry manure in Poland. Environ Sci Pollut Res Int. 2024;31(35):47727–41.

36. Wong MH, Chan EW, Chen S. IS26-mediated formation of a virulence and resistance plasmid in Salmonella Enteritidis. J Antimicrob Chemother. 2017;72(10):2750–4.

37. Hiley L, Fang NX, Micalizzi GR, Bates J. Distribution of Gifsy-3 and of variants of ST64B and Gifsy-1 prophages amongst Salmonella enterica Serovar Typhimurium isolates: evidence that combinations of prophages promote clonality. PLoS One. 2014;9(1):e86203.

38. Colavecchio A, D’Souza Y, Tompkins E, Jeukens J, Freschi L, Emond-Rheault JG, et al. Prophage Integrase Typing Is a Useful Indicator of Genomic Diversity in Salmonella enterica. Front Microbiol. 2017;8:1283.

39. Olaitan AO, Dia NM, Gautret P, Benkouiten S, Belhouchat K, Drali T, et al. Acquisition of extended-spectrum cephalosporin- and colistin-resistant Salmonella enterica subsp. enterica serotype Newport by pilgrims during Hajj. Int J Antimicrob Agents. 2015;45(6):600–4.

40. Han X, Peng J, Li J, Kuang R, Liu H, Xia J, et al. Genomic characterization of the convergence of multidrug resistance and virulence in Salmonella Enteritidis from a broiler slaughterhouse in China. Int J Food Microbiol. 2026;459:111920.

41. Li Y, Kang X, Ed-Dra A, Zhou X, Jia C, Muller A, et al. Genome-Based Assessment of Antimicrobial Resistance and Virulence Potential of Isolates of Non-Pullorum/Gallinarum Salmonella Serovars Recovered from Dead Poultry in China. Microbiol Spectr. 2022;10(4):e0096522.

42. Aung KT, Khor WC, Ong KH, Tan WL, Wong ZN, Oh JQ, et al. Characterisation of Salmonella Enteritidis ST11 and ST1925 Associated with Human Intestinal and Extra-Intestinal Infections in Singapore. Int J Environ Res Public Health. 2022;19(9).

43. Oladeinde A, Chung T, Bradshaw D, Diarra MS, Rehman A, Cook K, et al. The fitness of Salmonella Enteritidis in poultry is strongly shaped by interactions between bacteriophages, plasmids, and the microbial community. ASM Animal Microbiology. 2026;1(3).

44. Bolger AM, Lohse M, Usadel B. Trimmomatic: a flexible trimmer for Illumina sequence data. Bioinformatics. 2014;30(15):2114–20.

45. Wick RR, Judd LM, Gorrie CL, Holt KE. Unicycler: Resolving bacterial genome assemblies from short and long sequencing reads. PLoS Comput Biol. 2017;13(6):e1005595.

46. Schwengers O, Jelonek L, Dieckmann MA, Beyvers S, Blom J, Goesmann A. Bakta: rapid and standardized annotation of bacterial genomes via alignment-free sequence identification. Microb Genom. 2021;7(11).

47. Gurevich A, Saveliev V, Vyahhi N, Tesler G. QUAST: quality assessment tool for genome assemblies. Bioinformatics. 2013;29(8):1072–5.

48. Feldgarden M, Brover V, Gonzalez-Escalona N, Frye JG, Haendiges J, Haft DH, et al. AMRFinderPlus and the Reference Gene Catalog facilitate examination of the genomic links among antimicrobial resistance, stress response, and virulence. Sci Rep. 2021;11(1):12728.

49. Tian R, Zhou J, Imanian B. PlasmidHunter: accurate and fast prediction of plasmid sequences using gene content profile and machine learning. Brief Bioinform. 2024;25(4).

50. Carattoli A, Zankari E, Garcia-Fernandez A, Voldby Larsen M, Lund O, Villa L, et al. In silico detection and typing of plasmids using PlasmidFinder and plasmid multilocus sequence typing. Antimicrob Agents Chemother. 2014;58(7):3895–903.

51. Bharat A, Petkau A, Avery BP, Chen JC, Folster JP, Carson CA, et al. Correlation between Phenotypic and In Silico Detection of Antimicrobial Resistance in Salmonella enterica in Canada Using Staramr. Microorganisms. 2022;10(2).

52. Paradis E, Schliep K. ape 5.0: an environment for modern phylogenetics and evolutionary analyses in R. Bioinformatics. 2019;35(3):526–8.

53. Xu S, Li L, Luo X, Chen M, Tang W, Zhan L, et al. Ggtree: A serialized data object for visualization of a phylogenetic tree and annotation data. iMeta. 2022;1(4):e56.

54. Tonkin-Hill G, Lees JA, Bentley SD, Frost SDW, Corander J. RhierBAPS: An R implementation of the population clustering algorithm hierBAPS. Wellcome Open Res. 2018;3:93.

55. Croucher NJ, Page AJ, Connor TR, Delaney AJ, Keane JA, Bentley SD, et al. Rapid phylogenetic analysis of large samples of recombinant bacterial whole genome sequences using Gubbins. Nucleic Acids Res. 2015;43(3):e15.

56. Suchard MA, Lemey P, Baele G, Ayres DL, Drummond AJ, Rambaut A. Bayesian phylogenetic and phylodynamic data integration using BEAST 1.10. Virus Evol. 2018;4(1):vey016.

57. Volz EM, Didelot X. Modeling the Growth and Decline of Pathogen Effective Population Size Provides Insight into Epidemic Dynamics and Drivers of Antimicrobial Resistance. Syst Biol. 2018;67(4):719–28.

58. Jombart T, Ahmed I. adegenet 1.3-1: new tools for the analysis of genome-wide SNP data. Bioinformatics. 2011;27(21):3070–1.

59. Mook P, Gardiner D, Verlander NQ, McCormick J, Usdin M, Crook P, et al. Operational burden of implementing Salmonella Enteritidis and Typhimurium cluster detection using whole genome sequencing surveillance data in England: a retrospective assessment. Epidemiol Infect. 2018;146(11):1452–60.

60. Csardi G, Nepusz T. The igraph software package for complex network research. InterJournal, Complex Systems. 2006;1695(5):1–9.

61. Wick RR, Judd LM, Gorrie CL, Holt KE. Completing bacterial genome assemblies with multiplex MinION sequencing. Microb Genom. 2017;3(10):e000132.

62. Wick RR, Schultz MB, Zobel J, Holt KE. Bandage: interactive visualization of de novo genome assemblies. Bioinformatics. 2015;31(20):3350–2.

63. Grant JR, Enns E, Marinier E, Mandal A, Herman EK, Chen CY, et al. Proksee: in-depth characterization and visualization of bacterial genomes. Nucleic Acids Res. 2023;51(W1):W484–W92.

64. Camacho C, Coulouris G, Avagyan V, Ma N, Papadopoulos J, Bealer K, et al. BLAST+: architecture and applications. BMC Bioinformatics. 2009;10:421.

65. Wishart DS, Han S, Saha S, Oler E, Peters H, Grant JR, et al. PHASTEST: faster than PHASTER, better than PHAST. Nucleic Acids Res. 2023;51(W1):W443–W50.

