## Supplementary Figures for "Population Genomics of *Salmonella* Enteritidis in Saudi Arabia Reveals Globally Circulating Food- and Human-Associated Lineages and Plasmid-Mediated Antimicrobial Resistance"

### Supplementary Figures and Tables

**Supplementary Table S1 (SupplumentalTableS1.csv):** Accession numbers and metadata for study isolates and their associated plasmids

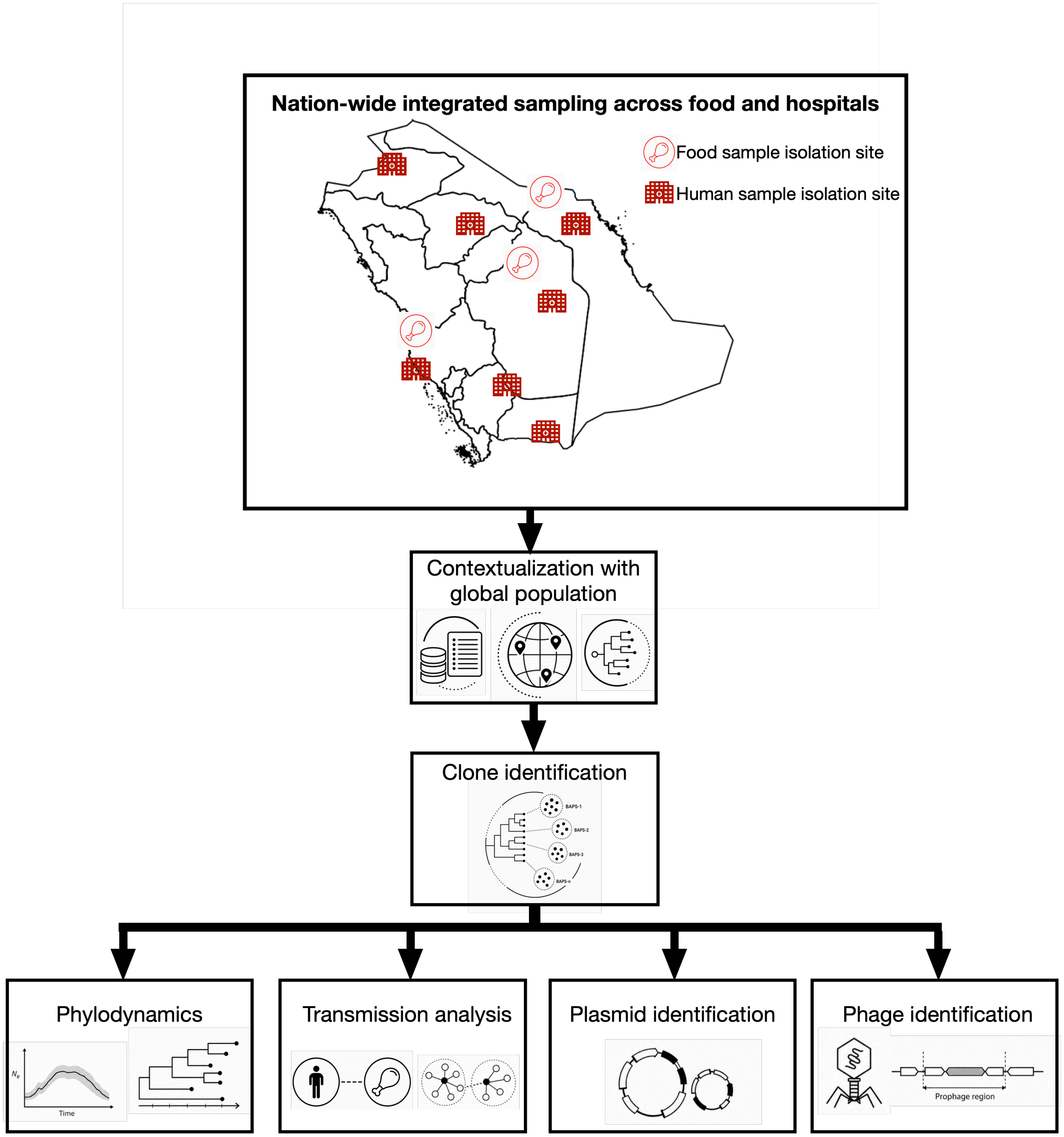


**Figure S1. Workflow of the study as detailed in Methods section.** The positions of icons on the map denote the isolation location. Details on metadata for isolates are provided in Supplemental Table S1.

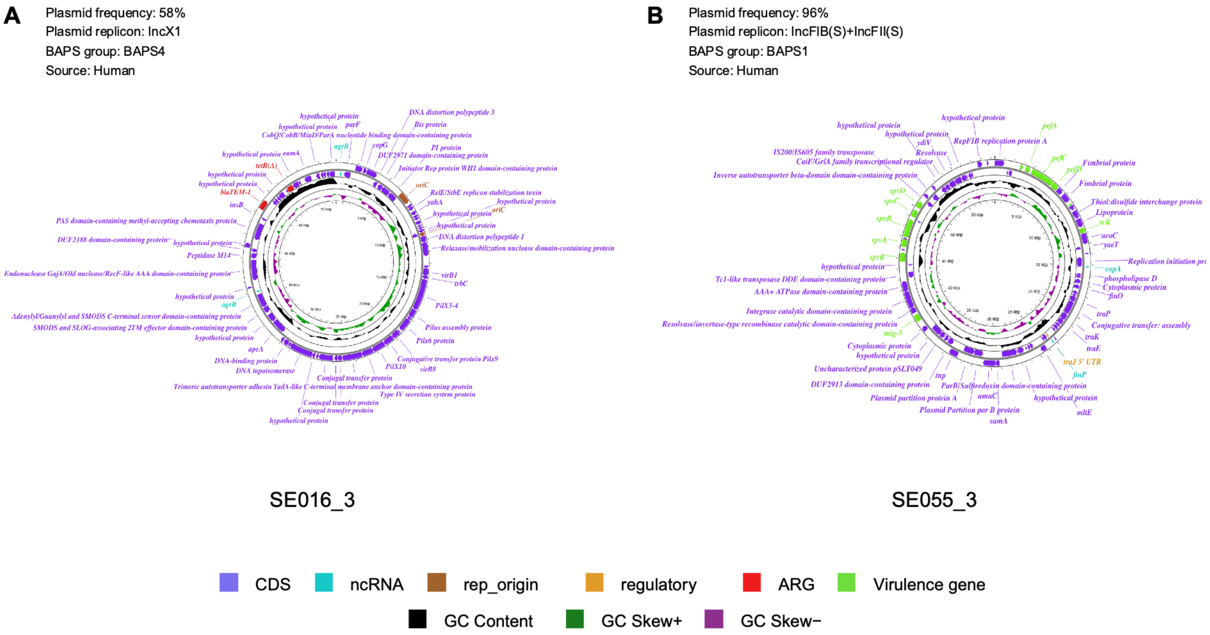


**Figure S2. Genomic maps of the two most frequently occurring *Salmonella* Enteritidis plasmids. (A)** A ~50-kbp resistance plasmid carrying bla$_{TEM-1}$ and *tetR(A)* together with a complete type IV secretion / conjugative-transfer system (*virB, pilX, tra*), indicating a self-transmissible resistance plasmid. **(B)** A ~60-kbp IncFIB(S)/IncFII(S) virulence plasmid carrying the *spv* operon (*spvABCDR*), the *pef* fimbrial cluster (*pefABCD*), *mig-5* and *rck*, with its own conjugative-transfer (*tra*) region. The phylogenetic distribution of these plasmids is shown in Figure 5. Source denotes the origin of the isolate from which the plasmid was extracted.

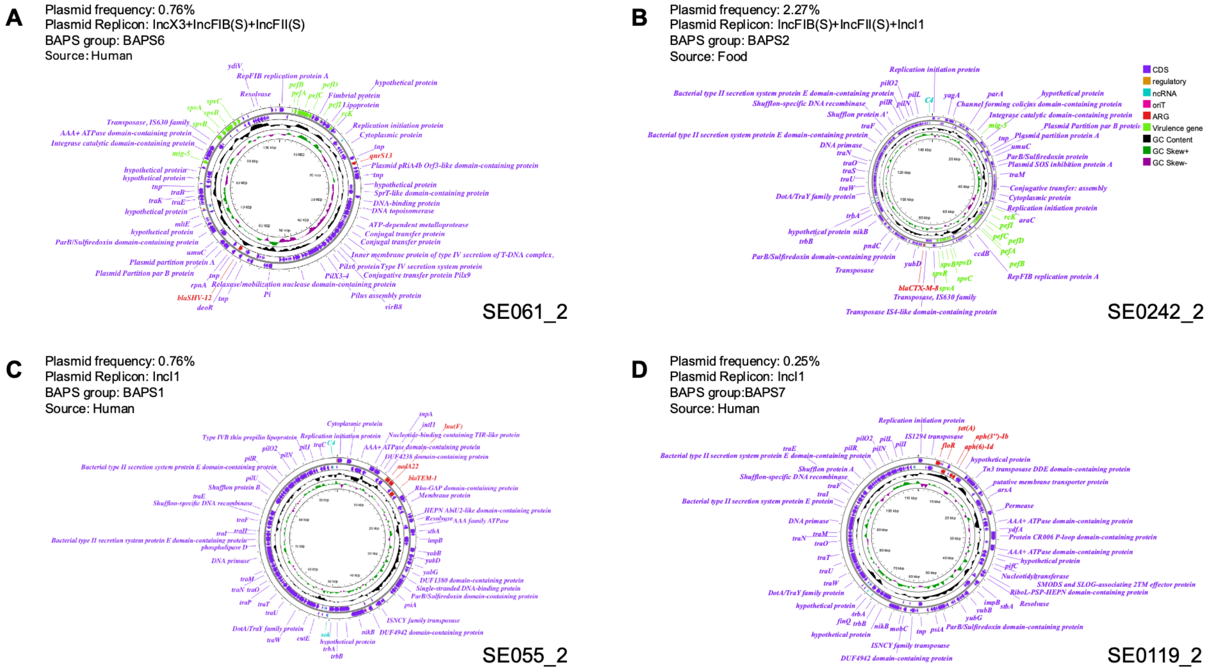


**Figure S3. Genomic maps of four extracted *S.* Enteritidis plasmids carrying antimicrobial resistance genes. (A)** SE061_2 (IncFIB, BAPS6, human), a hybrid virulence–resistance plasmid carrying *spv* and *pef* clusters together with *bla*$_{\text{SHV-12}}$ and *qnrS13*. **(B)** SE0242_2 (IncFIB, food), a hybrid virulence–resistance plasmid combining the *spv* operon (*spvABCDR*) and *pef* fimbrial cluster with *bla*$_{\text{CTX-M-8}}$. **(C)** SE055_2 (IncI1, BAPS 1, human), carrying *bla*$_{\text{TEM-1}}$, *aadA22* and *lnu(F)* alongside a complete *tra/pil* conjugative-transfer system. **(D)** SE0119, a resistance/conjugative plasmid carrying *tet(A), floR* and the aminoglycoside genes *aph(3'')-Ib / aph(6)-Id*, with multiple IS transposases and a type II secretion/conjugation system. The phylogenetic distribution of these plasmids is shown in Figure 5. Source denotes the source of the isolate, i.e. human or food, from which the plasmid was extracted.

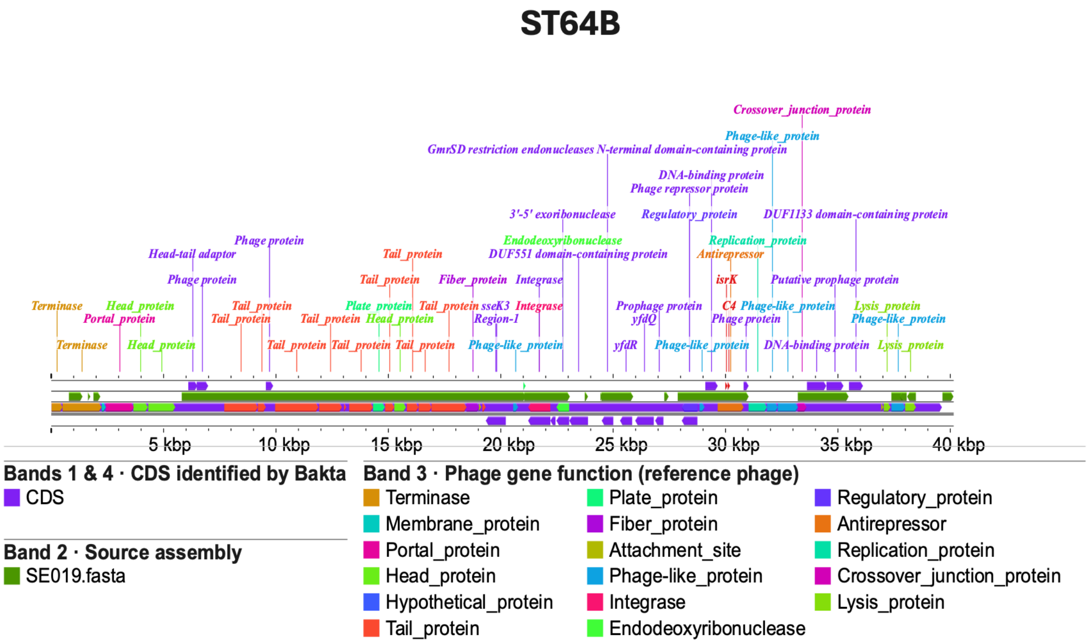


**Figure S4. Genomic map of the ST64B prophage identified in *S.* Enteritidis.** Prophage regions were predicted using PHASTEST, and genome annotation was performed using Bakta. The green band represents the alignment of a representative genome (SE019.fasta) against the ST64B prophage reference sequence using a 90% nucleotide identity threshold. The genomic map was generated using the built-in tools available in Proksee.
